# Business Models and Technologies of Leading Home Care Providers for Older Adults: A Multi-Country Comparative Study

**DOI:** 10.1101/2025.11.19.25340576

**Authors:** Panitda Huynh, Silvan Stöckli, Tobias Kowatsch, Rasita Vinay

**Author notes:** = shared last authorship (Tobias Kowatsch, Rasita Vinay).

## Abstract

**Background:** Global population ageing is accelerating due to declining fertility rates and rising life expectancy. Home care plays a central role in supporting older adults to remain in their own homes while managing ongoing care needs. However, home care provision is under mounting pressure, intensified by severe workforce shortages. Scalable, person-centred, and digitally supported home care services are therefore urgently required. This study addresses three questions: (RQ1) Who are the leading providers of home care for older adults? (RQ2) What business models do they employ? and (RQ3) What technologies do they use to deliver scalable care services?

**Methods:** This study used a mixed-methods, multiple case study design to investigate the business models of leading home care providers. To address RQ1, countries were identified using demographic and health system criteria, including the Global Innovation Index and population ageing indicators. Within each country, leading providers were selected through a purposeful, criterion-based sampling procedure informed by revenue, client volume, and expert validation. To address RQ2 and RQ3, we drew on four complementary data sources: (a) publicly available service descriptions, (b) provider information, (c) market and industry reports (including media and analyst coverage), and (d) semi-structured interviews with provider representatives and domain experts. Data were analysed using qualitative content analysis informed by the Business Model Canvas framework. A combined deductive and inductive coding approach was applied to identify business model patterns of technology integration across cases.

**Results:** This study examined leading home care providers in the most innovative countries Singapore, Switzerland, Sweden, the United Kingdom, and the United States, focusing on the three largest or most established providers in each country. Across the 15 cases, five recurring business model patterns were identified: (1) Franchise, (2) Fee-for-Service, (3) Diversified Customer Portfolio, (4) Community-based model, and (5) Performance-Based Contracting. Most providers adopted standard operational and clinical software (e.g., electronic medical records, scheduling, and billing systems). However, only a subset of providers - primarily in Singapore, Sweden, and the United States - implemented advanced digital solutions, such as predictive analytics, remote monitoring, robotic medication dispensers, and digital therapeutics.

**Conclusion:** This study provides the first cross-national comparison of home care business models and their use of digital health technologies, showing that digitalisation is driven mainly by operational efficiency. Scalable technology integration requires alignment between digital solutions, financing models, and ecosystem partnerships.

## 1. Introduction

The world is experiencing an unprecedented demographic shift as declining fertility rates and increasing life expectancy accelerate population ageing. By 2050, the proportion of adults aged 60 and older will more than double to 22% globally, intensifying chronic disease prevalence, functional decline, and care dependency (1). In OECD (Organisation for Economic Co-operation and Development) countries, public expenditure on long-term care is projected to rise above 3% of the GDP by 2050 (2). In response, many governments promote home care as a cost-effective alternative to institutional care, enabling older adults to age in place and maintain independence (2,3). Older adults often prefer to age in place, and home care models enable them to maintain quality of life (4). Between 2011 and 2021, the share of long-term care recipients has increased from 67% to 69% (5).

Despite its potential, the home care sector faces major structural challenges. A persistent shortage of caregivers, driven by shrinking family sizes, increased women’s participation in the workforce, and a shift from multi-generational households to older adults living independently, has strained care delivery (6,7). Caregivers are increasingly expected to not only support daily living but also to deliver complex medical and emotional care (6,8). These pressures have forced providers to reconsider how services (i.e., the provider and delivery of medical, nursing, and daily living support in home care) are organized and sustained. Research on the underlying business models is essential to help providers learn from successful approaches and design more efficient, scalable, and digitally-enabled care solutions (9–11).

Against this background, there is a pressing need to better understand how home care provision can be efficiently organised and scaled. Business models provide a conceptual lens to analyse how providers create, deliver, and capture value (12,13). In home care, reorganizing services and adopting new technologies often requires changes in reimbursement rules, professional roles, and care coordination. Because these elements are shaped by regulation and involve multiple stakeholders, business model innovation tends to be gradual and complex (13–15). While prior research has explored public-private partnerships (16), sustainable care models (17), and digital platform strategies (18), the specific business models of home care providers serving older adults remain underexplored. This is striking given that for-profit and non-profit providers are often at the forefront of innovation in response to demographic pressures, workforce shortages, and technological change (10).

In this study, we use the term digital health technologies (DHTs) to refer broadly to digital tools that support clinical care, operational efficiency, monitoring, or patient engagement (19). Although tools such as telecare, remote monitoring, and AI-assisted scheduling are increasingly deployed (20–23), little is known about how their adoption reshapes the underlying business models of home care providers. In particular, empirical comparative evidence across national contexts is scarce (9,24,25).

Despite the rapid growth of home care, there is limited empirical evidence on who the leading providers are and how they shape the sector across countries. To address this shortcoming, we formulate the first research question *(RQ1): Who are the leading providers of home care for older adults?* Second, while business models are increasingly recognised as essential to understanding how providers create, deliver, and capture value, the specific patterns employed by home care providers remain underexplored. To this end, we formulate the second research question *(RQ2): What business models do leading providers of home care employ?* Third, although DHTs are widely promoted as enablers of scalable and person-centred care, little is known about how such technologies are integrated into the business models of home care providers. To address this gap, we formulate the third research question *(RQ3): What technologies do leading providers of home care use to deliver scalable care services?*

By examining leading home care providers in the top five innovative countries, this study contributes to the limited but growing body of evidence on business models in home care. The findings aim to provide actionable insights for practitioners and policymakers seeking to develop scalable, digitally enabled, and person-centred home care services.

## 2. Methods

This study employed a qualitative multiple-case study design to explore leading home care providers across five countries. To address RQ1, we applied a structured provider selection process. To address RQ2, we analysed service information, provider data, market reports, and expert interviews using qualitative content analysis informed by the Business Model Canvas (BMC). Finally, to address RQ3, we classified DHTs using an adapted framework from the Digital Therapeutics Alliance (19). Each step of the process is detailed in the following subsections.

### 2.1 Study Design

We adopted a qualitative multiple case study design from a health systems and strategic management perspective, which is well suited to explore complex and context-dependent phenomena such as business model innovation in care delivery (26,27). Multiple case study approaches are recommended because they enable in-depth exploration of organisational processes while allowing cross-case comparison to identify patterns and variations across contexts, thereby increasing analytic generalisability (28,29). In health services research, case study designs are increasingly used to examine business models, innovation processes, and digital transformation (11,30). Building on previous research, our study focused on home care providers, combining documentary analysis, market reports, and semi-structured expert interviews (see Table 1). A cross-country comparative approach was chosen to capture contextual differences in health system structures, financing, and digitalisation, which are central to understanding the heterogeneity of home care systems.

**Table 1.** Data Sources.

| Type | Source I | Source II | Source III | Source IV |
| --- | --- | --- | --- | --- |
| Sources | Digital platforms; (website, app, etc.); service brochures; informational documents; media releases | Reports; Annual reports; financial reports | Media coverage (news articles, TV releases, etc.); market analyses; research publications | Interviews with experts |
| Purpose | Obtain insights into providers' service offerings, such as value propositions, target customer segments, key partners, and more | Gain insights into providers' competitive strategies, marketing approaches, and financial health | Understand providers' market positions and identify exemplary providers through market analysis and research | Gather additional insights to fill in gaps and validate information obtained from other sources |

*Note.* All data sources contributed to answering all three research questions (RQ1-RQ3). Each source type provides complementary information used for triangulation across cases.

### 2.2 Identifying Leading Countries and Providers of Home Care (RQ1)

To address RQ1, we implemented a structured selection approach. First, we selected the countries by applying five health- and innovation-relevant criteria adapted from prior comparative research (31–33): (1) position in the Global Innovation Index, (2) percentage of the population aged 65 years and older, (3) average percentage of healthy life expectancy at age 65, (4) Healthcare Access and Quality Index score, and (5) underlying healthcare system archetype, and confirmed the selection through expert consultation.

Second, leading providers were defined as those with a significant market position. Providers were evaluated against four equally weighted indicators: (a) annual revenue, (b) number of clients or service licenses, and (c) extent of integration of DHTs. The identification of leading home care providers followed a purposeful, four-step sampling procedure. First, publicly available data were triangulated from multiple sources, including provider websites, service brochures, press releases, annual reports, financial filings, market research reports, and governmental or professional association databases (see Table 1). Second, revenue was prioritized as the primary indicator of market leadership, in line with established research practices (38,39). In cases where such data were unavailable, proxy measures such as the number of clients, employees, or service locations were used to approximate market prominence (9,40). Third, we validated the preliminary provider selection with individuals who have professional expertise in the home care sector (e.g., researchers, consultants, or healthcare professionals). These experts were contacted individually and asked to confirm whether the selected providers are recognised as major or influential providers within their national home care market. This step ensured that the final provider list was contextually appropriate and reflective of the leading providers in each country. Finally, country-level demographic and economic characteristics and data for each identified provider were consolidated into a unified dataset encompassing indicators of organizational size, scope, and market presence, with the key metrics used for each country. To ensure robustness, we applied pairwise comparisons and a forward–backward selection procedure, with rankings further validated against expert opinions, official registries, and publicly available provider reports. The top three ranked providers in each country were included, resulting in 15 providers.

### 2.3 Identifying Business Models of Leading Providers (RQ2)

To address RQ2, we examined the business models of the selected home care providers using four complementary data sources (Table 1). Data were collected between May 2024 and February 2025. Following Matthews et al. (11,34), we conducted a document analysis using: (a) public provider materials, including service descriptions, websites, and digital brochures, (b) organizational documents, such as annual reports, governance statements, and strategy publications, (c) industry and policy reports, including national home care reports and comparative system analyses, (d) media coverage, retrieved from the Factiva database (news, press releases, and sector publications). All documents were retrieved through provider websites, government portals, and Factiva searches using provider name.

Semi-structured interviews were conducted to clarify how services were organized and how providers structured their business models. Interviews were conducted by PH and SS using a predefined interview guide (Appendix A). The guide was developed based on the BMC to cover all nine components (35), and included additional questions on how and where DHTs were embedded within service delivery, with examples provided during the interview to ensure conceptual clarity.

Participants were purposively selected based on their seniority and involvement in strategic or operational decision-making. Recruitment proceeded by: (a) identifying management-level staff on provider websites, (b) contacting them via LinkedIn or professional email, and (c) inviting participation through a standard study invitation message (Appendix B).

For each provider, we sought to conduct at least one interview to validate and refine the document-based business model analysis. Where interviews were possible, they were conducted in English, lasted 30–60 minutes, and were held via videoconference. All interviews were audio-recorded, transcribed verbatim, anonymised, and analysed using ATLAS.ti. An ethics exemption was obtained from the Ethics Committee of the University of St. Gallen for this study. All participants provided a written consent to participate in this study. Interviewee information is presented in Table A, with interview references cited by number.

For providers where interviews could not be conducted, business models were reconstructed exclusively from document analysis, using the same coding structure to ensure consistent cross-case comparison. Data saturation was defined as the point at which no new business model components or structural variations emerged across providers within the same country. Thus, saturation was assessed within-country, rather than across all 15 cases.

Two researchers, PH and SS, independently conducted and coded all interview and document data. An initial codebook was developed directly from the nine BMC components and iteratively refined during analysis to incorporate inductive sub-codes emerging from the data. Deductive coding was guided by the BMC, which includes nine elements: value propositions, customer segments, channels, customer relationships, revenue streams, key resources, key activities, key partnerships, and cost structure. After each coding round, the researchers met to reconcile interpretations and reach consensus, thereby ensuring analytic consistency and reliability. To mitigate potential bias, the initial coding was performed independently and blind to each other’s interpretations. Researcher positionality, defined as the reflexive awareness of how one’s background, assumptions, and institutional context may shape data interpretation, was continuously examined throughout the study to minimise interpretive bias (36). Bias was further minimised through systematic triangulation across data sources and iterative consensus meetings that enhanced analytical rigour and transparency.

We employed a directed qualitative content analysis combining both deductive and inductive approaches (37,38). In parallel, inductive coding was used to capture emergent themes such as policy adaptation, workforce innovation, and digital enablement. These codes were developed iteratively through repeated engagement with the data, allowing new concepts to emerge, be refined, and consolidated as patterns became evident across countries. Analysis proceeded in two phases: within-case analysis involved extracting and organizing data into BMC templates, validated through interviews and official documents; cross-case analysis involved systematic comparison to identify recurring business models and context-specific variations.

### 2.4 Identifying Digital Health Technologies in Home Care (RQ3)

To address RQ3, we analysed which DHTs were integrated into care delivery across providers. We first extracted all references to DHTs from the document-based data sources (see Table 1). We then classified each DHT according to the Digital Therapeutics Alliance framework (19). While originally designed for DHTs, we applied it here due to the absence of a dedicated framework for digital care services. From the full Digital Therapeutics Alliance framework, six categories were relevant to the DHTs observed in our dataset: (a) Health System Operational Software (e.g., scheduling, billing, and workflow coordination systems), (b) Health System Clinical Software (e.g., electronic medical records and clinical decision support tools), (c) Health & Wellness applications supporting wellbeing and lifestyle management, (d) Patient Monitoring, (e) Care Support tools assisting daily living, medication adherence, or psychosocial support, and (f) Digital Therapeutics, defined as evidence-based software interventions delivering therapeutic outcomes (see Appendix C). The remaining categories (Digital Diagnostics and Non-Health System Digital Solutions) were not identified among the included providers and were therefore excluded.

To complement and validate document-based classification, the semi-structured interviews included a dedicated set of questions on DHT use, asking participants to describe which DHTs were used and how they supported care delivery. Interview segments relating to DHTs were coded separately from business model components to maintain analytic distinction.

## 3. Results

We first present the leading home care providers across five countries and summarise the data sources included. Then, we outline the main business model patterns that emerged from the cross-case analysis and highlight contextual variations. Finally, we describe the range of DHTs used by providers and present differences in adoption and diversity across countries.

### 3.1 Identifying Leading Home Care Providers (RQ1)

#### 3.1.1 Country Identification

Singapore, Sweden, Switzerland, the United Kingdom, and the United States were selected based on predefined criteria reflecting demographic, economic, and innovation-related characteristics. Together, these indicators ensured a balanced representation of diverse health system models and stages of digital innovation. An overview of the comparative metrics across the five countries is presented in Table 2.

**Table 2.**
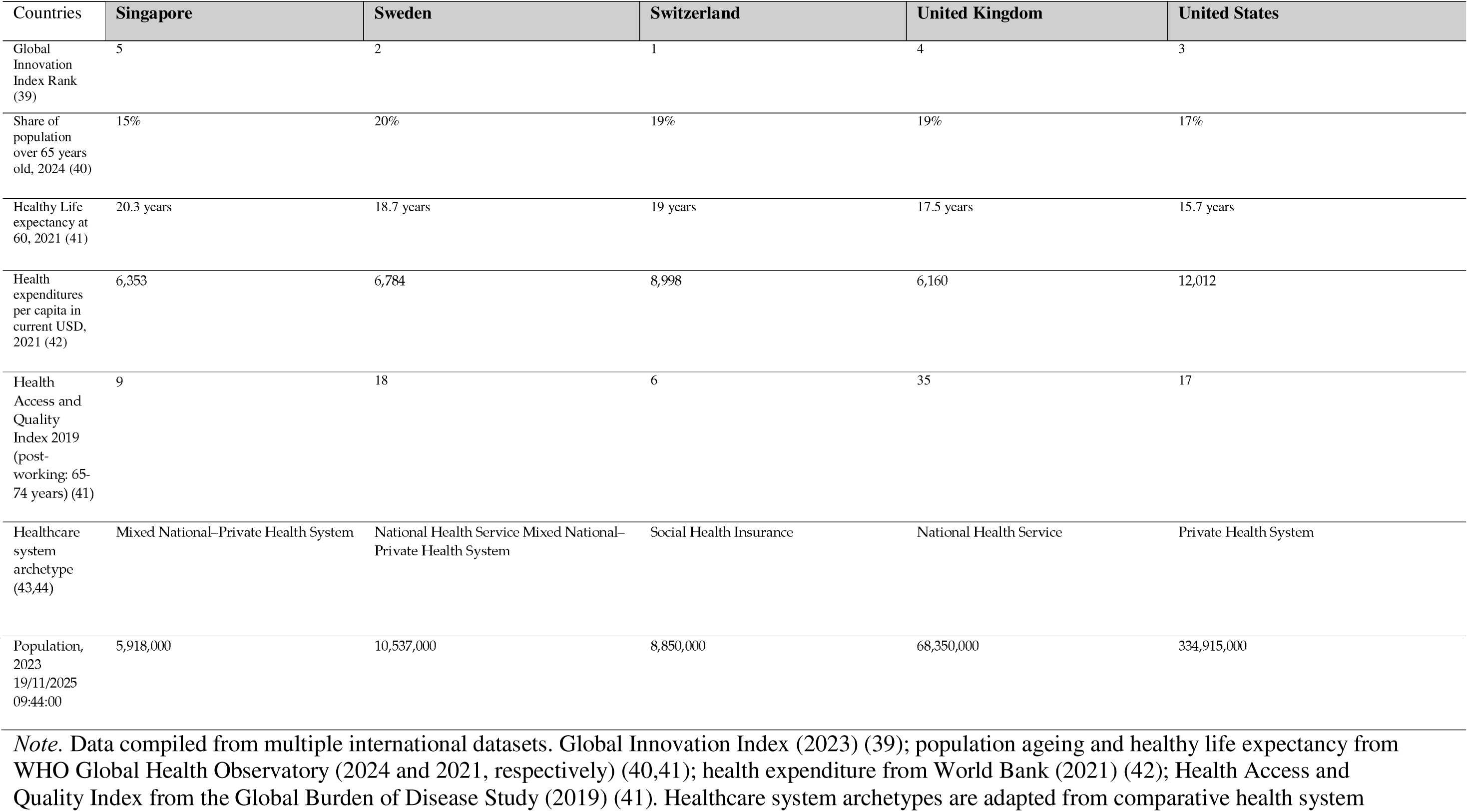
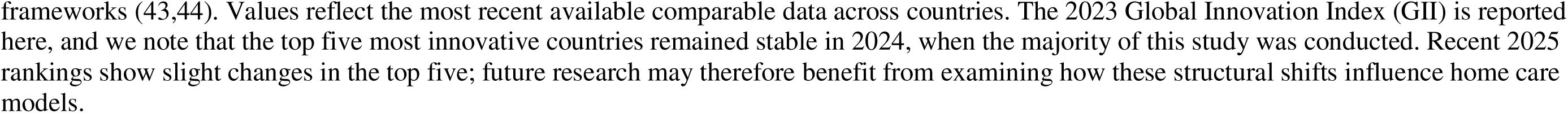
Overview of Selected Countries.

| Countries | Singapore | Sweden | Switzerland | United Kingdom | United States |
| --- | --- | --- | --- | --- | --- |
| Global Innovation Index Rank (39) | 5 | 2 | 1 | 4 | 3 |
| Share of population over 65 years old, 2024 (40) | 15% | 20% | 19% | 19% | 17% |
| Healthy Life expectancy at 60, 2021 (41) | 20.3 years | 18.7 years | 19 years | 17.5 years | 15.7 years |
| Health expenditures per capita in current USD, 2021 (42) | 6,353 | 6,784 | 8,998 | 6,160 | 12,012 |
| Health Access and Quality Index 2019 (post-working: 65-74 years) (41) | 9 | 18 | 6 | 35 | 17 |
| Healthcare system archetype (43,44) | Mixed National–Private Health System | National Health Service Mixed National–Private Health System | Social Health Insurance | National Health Service | Private Health System |
| Population, 2023<br>19/11/2025 09:44:00 | 5,918,000 | 10,537,000 | 8,850,000 | 68,350,000 | 334,915,000 |

*Note.* Data compiled from multiple international datasets. Global Innovation Index (2023) (39); population ageing and healthy life expectancy from WHO Global Health Observatory (2024 and 2021, respectively) (40,41); health expenditure from World Bank (2021) (42); Health Access and Quality Index from the Global Burden of Disease Study (2019) (41). Healthcare system archetypes are adapted from comparative health system
frameworks (43,44). Values reflect the most recent available comparable data across countries. The 2023 Global Innovation Index (GII) is reported here, and we note that the top five most innovative countries remained stable in 2024, when the majority of this study was conducted. Recent 2025 rankings show slight changes in the top five; future research may therefore benefit from examining how these structural shifts influence home care models.

#### 3.1.2 Provider Selection

Country-level demographic and economic indicators, together with provider-specific data, were consolidated into a unified dataset capturing measures of size, scope, and market presence. The key metrics applied for each country are summarised in Tables 2 and 3. Table 3 presents an overview of the leading home care providers across the five countries, demonstrating substantial variation in organisational scale, ownership structure, and market concentration. The data reveal a wide spectrum ranging from single-location non-profit providers in Singapore to large multi-branch enterprises such as Amedisys and Enhabit in the United States, reflecting the heterogeneous nature of home care markets internationally.

**Table 3.** Overview of Leading Home Care Providers.

| Providers | Singapore |  |  | Sweden |  |  | Switzerland |  |  | United Kingdom |  |  | United States |  |  |
| --- | --- | --- | --- | --- | --- | --- | --- | --- | --- | --- | --- | --- | --- | --- | --- |
|  | Home Nursing Foundation | St Luke’s Eldercare Ltd | All Saints Silver Lifestyle Club | Attendo Sverige AB | Vardaga Nytida Omsorg AB | Förenade Care AB | Institution genevoise de maintien à domicile (imad) | Spitex Zürich AG | Association pour la santé, la prévention et le maintien à domicile (ASPMAD) | Home Instead | Helping Hands | Bluebird Care | Maxim Healthcare Services | Amedisys Inc | Enhabit Home Health & Hospice |
| Revenue USD | 15,771,429 | 65,653,333 | 43,560,952 | 756,521,739 | 524,256,293 | 242,415,730 | 199,358,938 | 70,334,336 | 64,793,894 | 20,569,023 | 207,169,333 | 16,028,899 | 1,662,328,297 | 1,394,605,713 | 802,576,145 |
| Number of clients | 5,712 (only home care) | 1,206 | 1,060 | 5,907 | 2,147 | 1,387 | 14,856 | 6,735 | 5,055 | NA | NA | NA | NA | NA | NA |
| Number of Service Delivery Sites | 1 | 1 | 1 | 66 | 13 | 11 | 29 | 21 | 49 | 169 | 157 | 156 | 139 | 344 | 252 |

*Note.* Service delivery sites refer to branch offices, regional service hubs, or administrative units from which home care services are coordinated. This measure provides an indication of organisational scale rather than facility count.

### 3.2 Business Models of Home Care Providers (RQ2)

This study generated a comprehensive dataset comprising 159 documents and 11 semi-structured interview transcripts across four data modes from 15 providers (see Appendix D). Most documents (n=92) detailed service offerings, while 26 focused on provider-specific information and 30 on market reports. Interview data and document triangulation formed the basis for the comparative analysis of 15 home care providers across five countries. Table A provides an overview of participant roles and organizational contexts. A full overview of business model details is summarized in Appendix E.

Analysis across 15 providers revealed five recurring business model patterns: (1) Franchise, (2) Fee-for-Service, (3) Diversified Customer Portfolio, (4) Community-based model, and (5) Performance-Based Contracting. Together, these patterns illustrate the different strategic priorities providers adopt when seeking to balance scalability, financial sustainability, and responsiveness to care needs. The following subsections compare these patterns and their extensions.

**Table A.** Overview of Interviews.

| Participant code | Country | Participant characteristics | Duration (min) |
| --- | --- | --- | --- |
| P01 | Switzerland | CEO at a leading home provider; interviewed as expert | 60 |
| P02 | Switzerland | Head of Healthcare Services; interviewed as expert | 45 |
| P03 | United Kingdom | Director of Corporate Projects at a national home care provider; interviewed as provider representative | 45 |
| P04 | Switzerland | Lead Nurse and Project Quality Manager; interviewed as expert | 65 |
| P05 | Sweden | EVP, Strategy, Business Development & Quality; interviewed as provider representative | 55 |
| P06 | United Kingdom | Research Director specialising in long-term care markets; interviewed as expert | 40 |
| P07 | Switzerland | CIO at a home care provider; interviewed as provider representative | 40 |
| P08 | Switzerland | CEO of a regional home care provider; interviewed as provider representative | 30 |
| P09 | Switzerland | Owner and Manager of a private home care service; interviewed as provider representative | 64 |
| P10 | Singapore | Senior Researcher in ageing and care systems; interviewed as expert | 45 |
| P11 | Sweden | Chief Operational Excellence Officer at a large home care provider; interviewed as provider representative | 39 |

*Note.* P = Participant code; CEO = Chief Executive Officer; EVP = Executive Vice President; CIO = Chief Information Officer. Participant codes correspond to anonymised identifiers used in the qualitative analysis.

#### 3.2.1. Franchise-Based Model

Bluebird Care and Home Instead operate under a franchise model, a structure markedly distinct from direct service provision. In this model, the franchisor licenses its brand identity, operational systems, and training/support infrastructure to independent franchisees who are responsible for care delivery in designated territories.

Unlike traditional providers, the core value proposition for franchisors lies not in the execution of care services, but in the standardization and scaling of service models. This shifts the cost structure away from direct staffing and operational expenses toward support services, brand development, and quality control. The franchisors maintain brand integrity and regulatory alignment through structured audits, centralized training platforms, and shared digital infrastructures. Interview insights reinforced this delineation. *P03* noted that (see Table A):

> *“We are part of a franchise, which means we have to follow certain standards and protocols, but we also have the autonomy to adapt to our local market. That’s one of the strengths of the model - we get the support, the branding, and the system, but we can also innovate locally.” - (P03)*

Key elements enabling replication across franchise locations included standardized manuals, centralized electronic medical records (EMRs), and structured peer learning systems - features consistently emphasized by providers operating under the franchise model.

While this model was less prevalent in our global sample, it enables rapid geographic expansion without the capital intensity typically associated with opening new branches. This is particularly advantageous in fragmented markets or where regulatory systems favour local entrepreneurship. For instance, both providers demonstrated high regional penetration within the UK, with Bluebird Care operating over 200 franchises.

A key advantage of the franchise model lies in its dual adaptability: while the brand and core processes remain centralized, service delivery is locally customized. This balance of standardization and localization supports operational agility and strong customer rapport - both crucial in home care. However, this model also poses challenges in ensuring consistent care quality, especially as franchisees scale operations independently. Unlike vertically integrated providers, franchise-based systems rely heavily on the motivations and capabilities of individual owners, potentially leading to variability in service outcomes. Thus, the franchise model represents a distinctive and scalable pathway, aligning commercial replication strategies with localized home care delivery - a pattern that stands apart from the fee-for-service and government-contracted models seen elsewhere in this study.

#### 3.2.2. Fee-for-Service Model: Base Model

With the exception of the two franchisors, all other providers adopted a Fee-for-Service business model. In this pattern, services are clearly defined and delivered directly to clients in exchange for payment (see Figure 1). This approach is highly consistent across the analysed countries, albeit adapted to local reimbursement structures. Out-of-pocket payments remain common, though country-specific public co-funding mechanisms, health insurance schemes, and managed care arrangements influence the payment mix.

**Figure 1.**
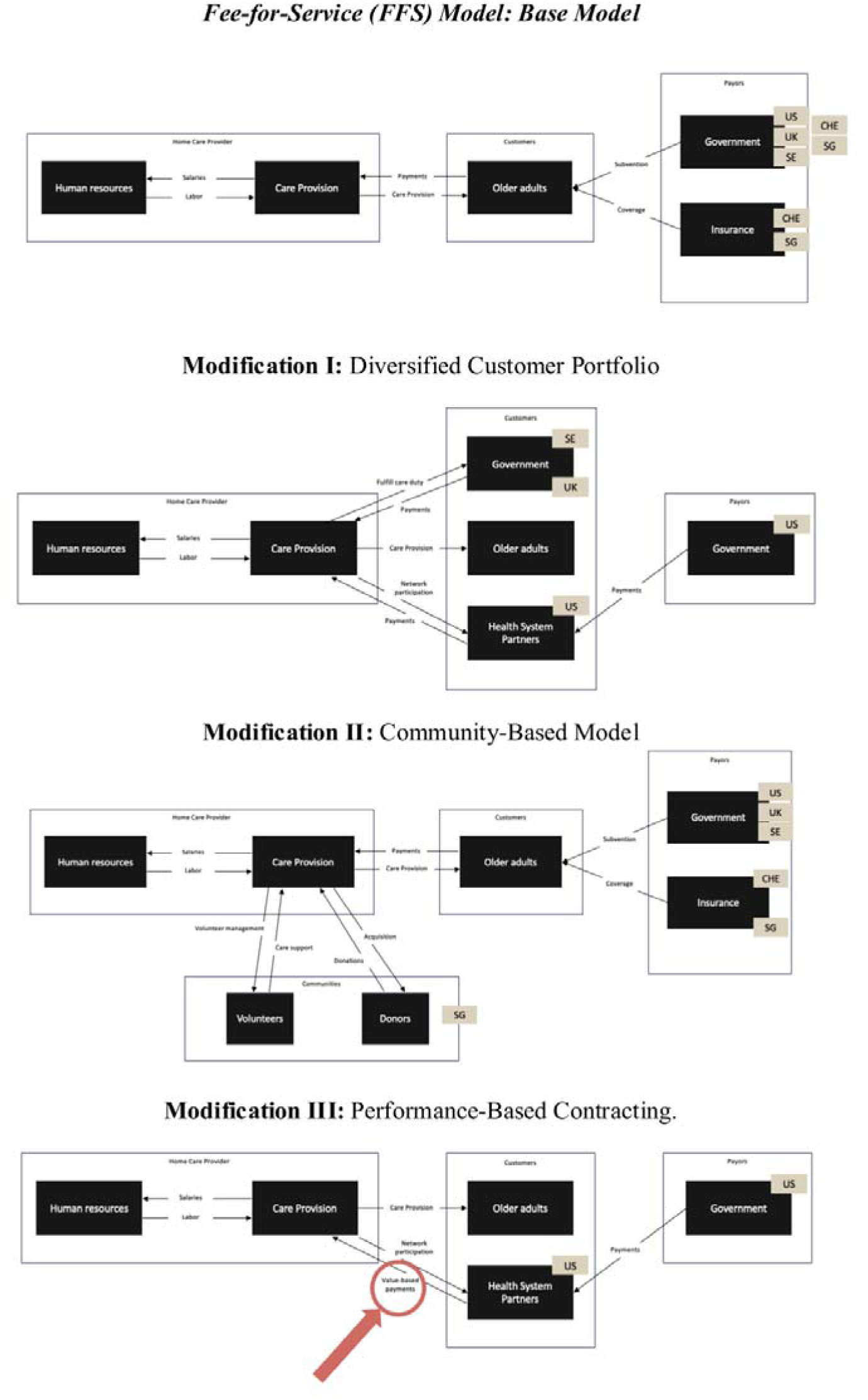
Overview of the Fee-for-Service Model and Its Modifications in Home Care. ***Note.*** The baseline fee-for-service model (top) ties revenue directly to the volume of care delivered. Modification I reflects diversification across B2C, B2G, and B2B payer relationships. Modification II incorporates philanthropic funding, volunteer support, and community partnerships to expand service scope. Modification III introduces performance-linked reimbursement elements, aligning revenue with quality and outcomes. Arrows indicate the direction of value, service, and data flows between business model components.

Interview evidence reinforced this pattern. For example, P05 confirmed: *“Our model is mainly based on individual care plans where the municipality pays per hour of delivered service. It’s very transparent”.* Similarly, P08 described the billing structure in Switzerland: *“We have to follow tariff agreements, so the services are priced and invoiced by the minute, and there’s a strong regulation from insurers and the canton.”*

Revenue streams are therefore largely tied to the actual volume of services delivered - whether to municipalities, insurers, or directly to clients. Staffing remains the primary cost driver, especially as wages often account for over 80% of operational costs in home care. The high labour-intensity of the model also constrains scalability and profitability, as echoed by stakeholders in Sweden and the UK. This is a sample quote (P11): *“Everything revolves around having the right carer at the right time. That’s where most of the cost and coordination lies.”*

While operational frameworks vary in terms of scheduling sophistication and resource allocation tools, the foundational logic of exchanging service time for fees remains unchanged. Providers typically manage a balance between efficiency and personal care, often using route optimization and case management tools to contain costs while maintaining care quality.

Overall, the Fee-for-Service model represents the dominant pattern due to its compatibility with existing funding systems and regulatory frameworks. Its pervasiveness stems from its straightforward alignment with traditional healthcare accounting and public procurement procedures. However, its sustainability is increasingly questioned considering demographic pressures and workforce shortages.

Across providers, the Fee-for-Service model represents the baseline business model logic, in which revenues are tied to the volume of care delivered. From this baseline, we observed three distinct modifications that adapt the core model to different strategic priorities: (3) Diversified Customer Portfolio, (4) Community-Based, and (5) Performance-Based Contracting. The following subsections elaborate on these extensions and the conditions under which they emerge.

#### 3.2.3 Variants of the Fee-for-Service Model

##### 3.2.3.1 Modification I: Diversified Customer Portfolio

A prominent modification observed across several providers was the strategic diversification of customer segments beyond traditional business-to-consumer (B2C) relationships. This evolution toward multi-sided market patterns reflects a context-sensitive response to national funding structures and policy environments. In Sweden, for instance, providers such as Attendo and Ambea concurrently operate under both business-to-government (B2G) and B2C arrangements, capitalizing on the statutory role of municipalities in financing home care.

In the United States, providers extend their revenue base through business-to-business (B2B) partnerships, particularly with Medicare Advantage and managed care organizations. Medicare Advantage refers to private health plans that administer Medicare benefits under contract with the U.S. federal government, reimbursing providers for covered services delivered to enrolled beneficiaries (45). Under these arrangements, providers contract directly with insurers or care management entities to deliver home-based personal or skilled nursing services reimbursed on a fee-for-service basis. Such partnerships enable providers to access a broader pool of insured clients while ensuring steady reimbursement flows within the existing payment framework (46,47). By contrast, UK providers explained that, although they qualified to participate in local government commissioning frameworks, they chose not to do so because of misaligned financial incentives and high administrative burden.

These findings illustrate a key variation within the fee-for-service model: a deliberate expansion of customer interface strategies across B2C, B2G, and B2B segments. This strategic adaptation enables providers to buffer financial volatility, meet contractual diversity, and navigate the regulatory heterogeneity across national contexts. The structural logic of these adaptations is illustrated in Figure 1.

##### 3.2.3.2 Modification II: Community-Based Model

In Singapore, fee-for-service models were notably augmented through deep-rooted community engagement, particularly among non-profit providers. These organizations strategically leveraged philanthropic support and volunteerism to broaden their service scope beyond core reimbursable care. While foundational medical and nursing services were financed through public subsidies or insurance schemes, supplementary offerings - such as home modifications, caregiver training, and social participation programs - were funded through charitable donations and community partnerships. Another sample quote (P10) from the interview stated: “*For non-profits, they are very much relying on donations to provide the additional service. not necessarily basic medical care, but it could be things like home modification, assistive devices, or caregiver support services*.”

This model extension reflects a hybrid funding approach wherein community-driven contributions complement rather than substitute formal payment structures. It enables a more holistic response to aging-in-place needs, tailored to socio-cultural expectations and resource limitations. In contrast, Swiss non-profit providers, despite operating under a similar organizational status, indicated minimal reliance on philanthropic inputs, citing stronger statutory funding channels and a more regulated reimbursement environment. Figure 1 depicts this differentiated model pattern, highlighting the extent to which community involvement is structurally embedded within the broader financial architecture of service provision.

##### 3.2.3.3 Modification III: Performance-Based Contracting

A third variation of the fee-for-service model observed across several countries involved the integration of value-based payment (VBP) mechanisms. This emerging pattern introduces performance-linked reimbursement components into traditional service delivery, aligning provider incentives with health outcomes, care quality, and client satisfaction rather than service volume alone. These arrangements involve contractual agreements in which providers deliver home-based services reimbursed under capitated or value-based payment models, thereby securing more stable and diversified revenue streams (45). While still rooted in fee-for-service logic, VBP frameworks embed contractual expectations regarding efficiency, effectiveness, and user-centred metrics.

In the United States, two providers reported active participation in Medicare’s Home Health Value-Based Purchasing model, which directly ties reimbursement to provider performance on quality indicators such as hospital readmission rates, client satisfaction, and care coordination. This strategic adaptation reflects a fundamental shift in revenue logic from volume-based compensation toward outcomes-oriented incentives. Organizational transformation to meet these benchmarks was often substantial, involving investments in data infrastructure, integration with EMRs, and the promotion of inter-professional collaboration.

Similarly, Swedish providers operating under municipal contracts described initial efforts to incorporate quality-linked components into their reimbursement logic, though implementation remained limited. These accounts highlight the gradual evolution of public contracting environments toward blended models that reward both service provision and measurable outcomes.

Overall, these emergent arrangements suggest a transitional state between traditional fee-for-service and fully outcome-based contracting. Although not yet standardized or widespread, their presence across innovation-forward systems signals a growing convergence with broader health system goals emphasizing value over volume. Figure 1 illustrates this hybrid configuration, capturing the blended logic of quantifiable service delivery and performance-based incentives.

### 3.4 Digital Health Technologies Used in Home Care (RQ3)

The third research question explored DHTs that home care providers integrated into care. Across the 15 providers, a total of 45 distinct DHTs were identified (Table 4), categorized into six domains according to Digital Therapeutics Alliance - which are: Health & Wellness, Monitoring, Care Support, Digital Diagnostics, Digital Therapeutics, Health System Clinical Software, Health System Operational Software, and Non-Health System Software / Digital Health Solutions (19). The majority were classified under Health System Clinical Software (n=17; 38%) and Health System Operational Software (n=14; 31%), underscoring a strategic emphasis on clinical documentation, care coordination, and internal efficiencies.

**Table 4.**
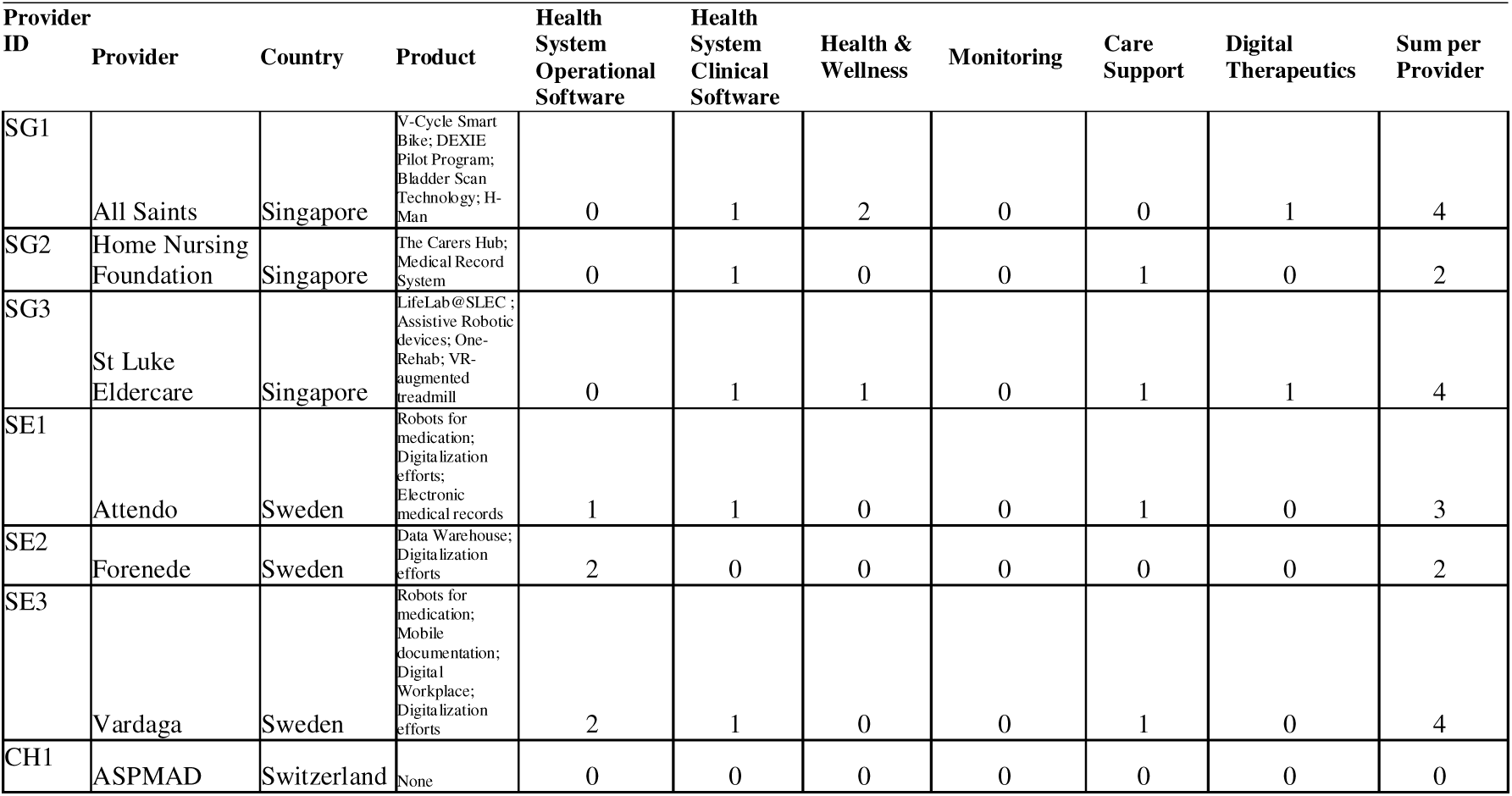
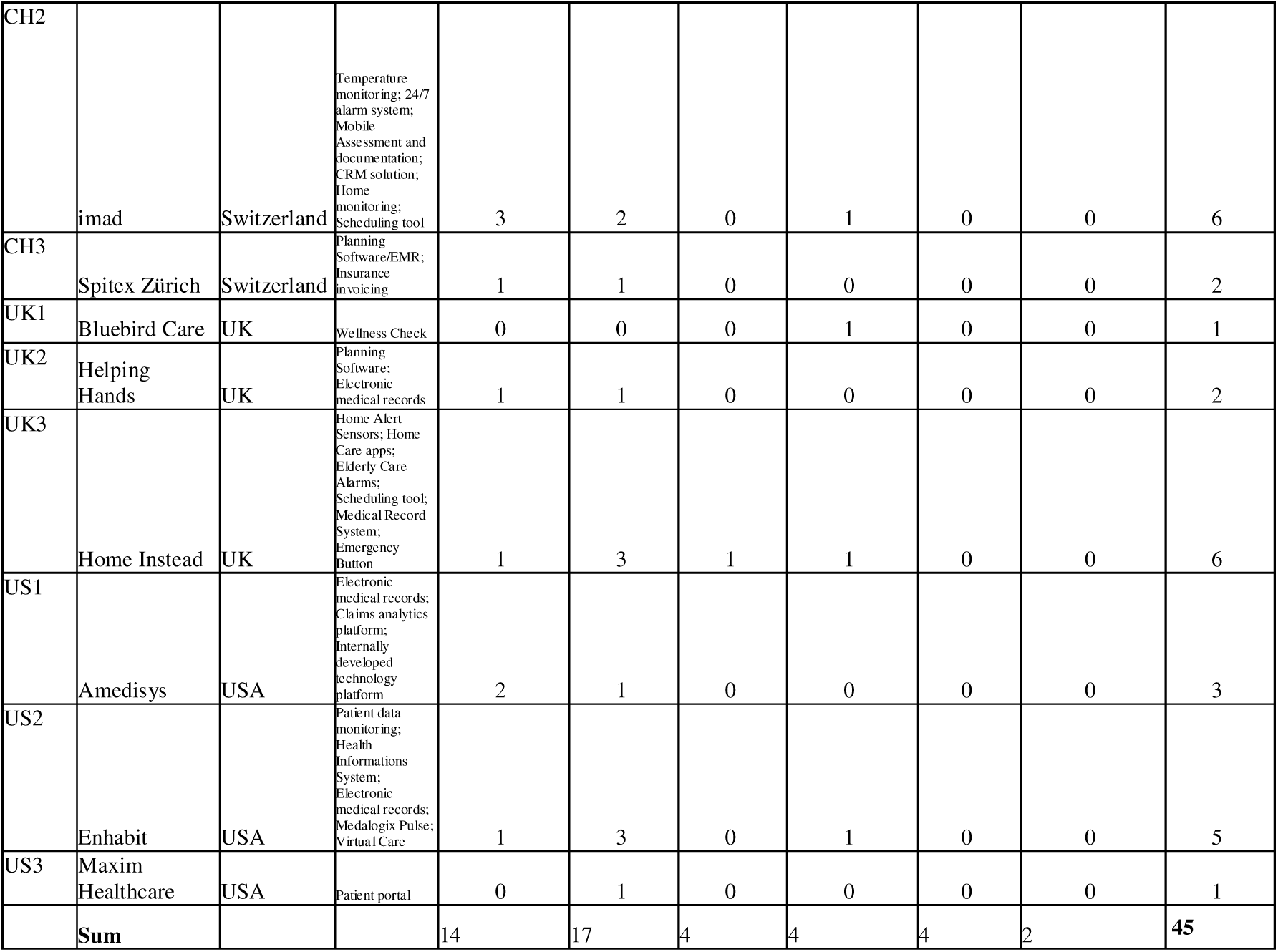
Identified Digital Health Technologies across Providers.

| Provider ID | Provider | Country | Product | Health System Operational Software | Health System Clinical Software | Health & Wellness | Monitoring | Care Support | Digital Therapeutics | Sum per Provider |
| --- | --- | --- | --- | --- | --- | --- | --- | --- | --- | --- |
| SG1 | All Saints | Singapore | V-Cycle Smart Bike; DEXIE Pilot Program; Bladder Scan Technology; H-Man | 0 | 1 | 2 | 0 | 0 | 1 | 4 |
| SG2 | Home Nursing Foundation | Singapore | The Carers Hub; Medical Record System | 0 | 1 | 0 | 0 | 1 | 0 | 2 |
| SG3 | St Luke Eldercare | Singapore | LifeLab@SLEC; Assistive Robotic devices; One-Rehab; VR-augmented treadmill | 0 | 1 | 1 | 0 | 1 | 1 | 4 |
| SE1 | Attendo | Sweden | Robots for medication; Digitalization efforts; Electronic medical records | 1 | 1 | 0 | 0 | 1 | 0 | 3 |
| SE2 | Forenede | Sweden | Data Warehouse; Digitalization efforts | 2 | 0 | 0 | 0 | 0 | 0 | 2 |
| SE3 | Vardaga | Sweden | Robots for medication; Mobile documentation; Digital Workplace; Digitalization efforts | 2 | 1 | 0 | 0 | 1 | 0 | 4 |
| CH1 | ASPMAD | Switzerland | None | 0 | 0 | 0 | 0 | 0 | 0 | 0 |
| CH2 | imad | Switzerland | Temperature monitoring; 24/7 alarm system; Mobile Assessment and documentation; CRM solution; Home monitoring; Scheduling tool | 3 | 2 | 0 | 1 | 0 | 0 | 6 |
| CH3 | Spitex Zürich | Switzerland | Planning Software/EMR; Insurance invoicing | 1 | 1 | 0 | 0 | 0 | 0 | 2 |
| UK1 | Bluebird Care | UK | Wellness Check | 0 | 0 | 0 | 1 | 0 | 0 | 1 |
| UK2 | Helping Hands | UK | Planning Software; Electronic medical records | 1 | 1 | 0 | 0 | 0 | 0 | 2 |
| UK3 | Home Instead | UK | Home Alert Sensors; Home Care apps; Elderly Care Alarms; Scheduling tool; Medical Record System; Emergency Button | 1 | 3 | 1 | 1 | 0 | 0 | 6 |
| US1 | Amedisys | USA | Electronic medical records; Claims analytics platform; Internally developed technology platform | 2 | 1 | 0 | 0 | 0 | 0 | 3 |
| US2 | Enhabit | USA | Patient data monitoring; Health Informations System; Electronic medical records; Medalogix Pulse; Virtual Care | 1 | 3 | 0 | 1 | 0 | 0 | 5 |
| US3 | Maxim Healthcare | USA | Patient portal | 0 | 1 | 0 | 0 | 0 | 0 | 1 |
|  | **Sum** |  |  | 14 | 17 | 4 | 4 | 4 | 2 | 45 |

#### 3.4.1 Health System Clinical Software

Clinical Software tools included EMRs used by nurses and care professionals for documentation and history tracking. Many systems featured front-end interfaces for older adults and family members, enabling transparency over delivered services and time allocation. Some platforms allowed for the purchase of additional services, and several incorporated clinical decision support with predictive analytics. Emergency-response technologies such as fall sensors, and wearable buttons were also found in this category. Fall sensors were classified under Health System Clinical Software rather than Patient Monitoring, because their primary function is to trigger real-time clinical or emergency intervention rather than to support continuous physiological or behavioural monitoring. While Patient Monitoring systems passively collect and analyse data trends (e.g., movement patterns, hydration levels, or vital signs) to inform preventive care, emergency-response technologies operate within the clinical workflow, activating alerts that connect directly to caregivers, clinical staff, or emergency services. In line with the Digital Therapeutics framework, DHTs that enable immediate clinician engagement or direct linkage to care delivery systems are therefore considered clinically integrated tools rather than passive monitoring devices. Moreover, telemedicine portals were also classified under Health System Clinical Software, as they support clinician-patient interactions and enable the provision of remote care.

#### 3.4.2 Health System Operational Software

Operational Software comprised route planning systems, staff scheduling tools, claims analytics platforms, and internal communication intranets. These technologies enabled efficiency gains and supported regulatory compliance, making them attractive in both public and private provider contexts.

#### 3.4.3 Patient Monitoring

Technologies within the Patient Monitoring domain (n=4; 8%) included home-based sensor systems to track hydration, movement, and environmental hazards. Analytics platforms enabled risk stratification and early warning score generation, facilitating preventative interventions.

#### 3.4.4 Health & Wellness

Health & Wellness technologies (n=4; 8%) included robotic fitness devices, cognitive gaming robots, and digitally enhanced stationary bikes. These technologies primarily supported functional maintenance and social participation rather than direct clinical outcomes.

#### 3.4.5 Care Support

Care Support technologies (n=4; 8%) were most prominent in Singapore and Sweden. In Singapore, providers deployed digital ecosystems offering educational resources, peer-to-peer support, and product marketplaces. On-site immersive experiences helped family members understand care routines. In Sweden, robotic medication dispensers tracked adherence and ensured timely administration, aligning with municipal quality mandates.

#### 3.4.6 Digital Therapeutics

Two technologies were categorised as Digital Therapeutics (n=2; 4%): the H-Man robotic rehabilitation device for upper-limb recovery and a VR-augmented treadmill designed to improve mobility and balance. Both were deployed in Singapore and reflect emerging trends toward function-focused therapeutic innovations. In this study, the Digital Therapeutics category was applied analytically following the Digital Therapeutics Alliance framework, encompassing software-driven interventions that produce measurable therapeutic effects, regardless of national reimbursement or regulatory status. While countries such as the United States and Singapore have advanced ecosystems supporting digital therapeutics innovation, others (e.g., Switzerland, Sweden, and the United Kingdom) lack formal reimbursement pathways or regulatory definitions comparable to those established in Germany. Consequently, technologies were classified according to their functional characteristics and intended clinical outcomes rather than their legal recognition within each national context.

#### 3.4.7 Cross-Country Variation in Digital Portfolios

Country-level variation was notable. Swiss and UK providers concentrated on EMRs and planning tools, reflecting operational and regulatory needs. U.S. providers showed more integration of predictive analytics and monitoring, aligned with value-based reimbursement. Sweden adopted internal data warehouses, digital transformation strategies, and robotic adherence tools. Singapore exhibited the highest technological diversity (n=10), including VR, robotics, and wellness-oriented technologies - many implemented onsite at care facilities rather than in individual homes. This breadth reflects Singapore’s integrated, facility-based care delivery model and strong public-private partnership culture. Conversely, DHTs categorized as Non-Health System Software and Digital Diagnostics were absent. This may be attributed to regulatory uncertainties, reimbursement limitations, or perceived misalignment with core home care services.

## 4. Discussion

In the following sections, we first discuss the practical implications of the findings. We then outline the theoretical implications, focusing on how the results extend current understandings of business model adaptation and digital transformation in home care. Next, we critically examine the limitations of the study in relation to research design, data availability, and generalisability. Finally, we propose directions for future research, highlighting areas that warrant deeper empirical and conceptual investigation.

### 4.1 Practical Implications

The findings of this study offer several important implications for practice. The identification of leading providers through a multidimensional strategy - combining revenue, client base, and geographic scope - moves beyond conventional single-metric rankings and demonstrates that organisational prominence in home care cannot be assessed solely through financial performance (10,11,34). Franchise-based providers in particular disrupted expected correlations between revenues and client base, revealing that scale and consolidation do not always align. For policymakers and regulators, this underscores the need to interrogate the structural underpinnings of organisational growth and to strengthen harmonised reporting standards that facilitate benchmarking and accountability across countries. Without more transparent and comparable metrics, cross-national policy learning and oversight will remain hampered.

The analysis of business model patterns further highlight the consequences of reimbursement and financing logics for long-term sustainability. Fee-for-Service arrangements, although administratively straightforward and widely entrenched, risk prioritising throughput over outcomes. In aging societies with rising multimorbidity and growing workforce shortages, this creates pressures that may undermine care quality and exacerbate strain on providers. The gradual emergence of hybrid models and value-based care elements, particularly in Sweden and the United States, illustrates that transitions are possible but resource-intensive, requiring substantial investments in digital infrastructure, workforce training, and data integration. Providers and funders should therefore anticipate that such transitions will not be uniform but will instead demand targeted support and adaptive regulatory frameworks. At the same time, diversification of customer segments - as observed in the United States, Sweden, and the UK - offers opportunities for financial resilience and cross-subsidisation, yet also raises questions about governance and equity. These tensions should be carefully managed by commissioners and payers to balance financial feasibility with service quality.

DHT adoption patterns similarly point to practical implications for innovation policy. The strong concentration of tools in the domains of Health System Clinical Software (41%) and Operational Software (30%) reflects a prevailing emphasis on compliance, efficiency, and cost containment. While such technologies are essential for documentation and billing, the underrepresentation of Monitoring, Health & Wellness, Care Support, and Digital Therapeutics categories reveals missed opportunities to strengthen prevention, engagement, and functional outcomes (24,25,48,49). Incentive structures, data interoperability, and training support must be strengthened to enable wider uptake of these more transformative tools. Cross-national differences further illustrate the decisive influence of systemic context: Singapore’s mix of funding sources and focus on results made it possible to test new digital therapies and wellness tools. In contrast, Switzerland and the UK took a more cautious approach, concentrating on improving day-to-day operations but held back by fragmented systems and older governance structures. Practically, this means that policy transfers should be pursued with caution, as organisational strategies are deeply conditioned by systemic incentives and cultural norms.

Finally, the classification and adoption of Digital Therapeutics warrant specific attention. A further consideration concerns the definition and recognition of Digital Therapeutics, which vary significantly across national contexts. While the Digital Therapeutics Alliance provides an internationally accepted functional definition - referring to software-based interventions that deliver evidence-based therapeutic outcomes - formal classification and reimbursement mechanisms remain uneven. For example, Germany has established a dedicated reimbursement pathway for digital therapeutics under the Digitale Gesundheitsanwendungen (DiGA) framework, whereas Switzerland and Sweden currently lack equivalent regulatory or payment structures. Consequently, our study applied the Digital Therapeutics category analytically based on the DHTs’ functional and therapeutic intent rather than their formal status within each health system. These discrepancies underscore that cross-country comparisons of digital therapeutics must consider regulatory maturity and reimbursement incentives, which critically shape providers’ adoption decisions and the scalability of such innovation.

### 4.2 Theoretical Implications

While existing literature often focuses on macro-level typologies or single delivery innovations (24,34,50,51), our analysis shows how providers actively reconfigure business models in response to local funding structures, regulation, and societal expectations. The persistence of Fee-for-Service as the dominant business model architecture, consistent with the Health@Home logic described in previous research (50), raises questions about the long-term sustainability of volume-based reimbursement in light of demographic and workforce pressures. Theoretically, this underscores the limits of efficiency-driven models that lack flexibility to address complex and evolving care needs. The evidence that Fee-for-Service models may encourage throughput rather than outcomes contributes to a growing literature warning against the unintended consequences of volume-oriented reimbursement systems in ageing societies.

The emergence of hybrid and modified patterns in the United States and Sweden provides further theoretical insight into how institutional environments condition business model innovation. The partial and uneven integration of value-based care logics illustrates that transitions are rarely linear, but instead involve a process of institutional negotiation and adaptation. Providers face the dual challenge of aligning with new performance-linked incentives while remaining compliant with legacy Fee-for-Service structures, a dynamic that highlights the concept of path dependency in healthcare business model evolution. Such dynamics demonstrate that institutional and resource constraints shape not only the pace but also the pattern of innovation, extending theories of hybridisation in health service delivery.

The diversification of customer segments observed in the U.S., Sweden, and the UK offers theoretical contributions to debates on multi-payer systems and organisational resilience. While diversification reduces exposure to reimbursement shocks and enables cross-subsidisation, it also raises governance and accountability concerns, particularly when public budgets are constrained. This points to a broader theoretical tension: business model flexibility may enhance organisational survival, but its impact on equity and service quality depends heavily on the distribution of bargaining power across institutional actors.

The outlier case of Singapore highlights the importance of theorising non-commercial forms of value capture. By embedding state subsidies within a socio-civic fabric of donations, volunteer labour, and philanthropic support, Singaporean providers institutionalise social capital and community legitimacy as integral components of their business models. This finding demonstrates that the BMC (52), while effective in structuring analysis, requires refinement to incorporate relational and civic value mechanisms that extend beyond conventional market-based categories.

The franchise model observed in the UK further enriches theoretical discussions on standardisation and adaptability. While franchising enables scalability and operational efficiency, it may constrain local responsiveness and innovation by prioritising brand consistency and compliance. This shows a balance between providers becoming more alike and their ability to adapt, which matters especially in fast-changing and unpredictable policy settings.

Finally, the analysis of contextual embeddedness across systems challenges the explanatory sufficiency of classical welfare regime typologies (24,53). Although countries such as Sweden and the UK are grouped under the Beveridge model, their home care sectors differ substantially in governance and market orientation. Similarly, Switzerland’s Bismarckian framework has hybridised through cantonal interventions, while Singapore’s model defies binary classification by combining mixed financing with strong state coordination and public-private partnerships. These findings suggest that macro-level welfare regime theory must be complemented by meso-level institutional and governance analysis to capture the diversity of provider strategies. They also highlight that business model components - such as value propositions, revenue streams, and partnerships - are highly sensitive to systemic context and not universally portable.

The findings on DHT adoption further extend theories of digital transformation in healthcare. Rather than following a uniformly disruptive trajectory, digitalisation in home care is shown to be compliance- and efficiency-driven, privileging administrative and regulatory imperatives over patient-facing innovation. This supports theoretical perspectives that emphasise the institutional conditioning of technology adoption, where regulatory mandates, reimbursement incentives, and vendor ecosystems shape trajectories more decisively than technological potential alone (18,30,54). Cross-national patterns reinforce this argument: Singapore’s outcome-oriented financing facilitated early experimentation with digital therapeutics, Sweden, and the U.S. integrated predictive analytics in line with value-based reforms, while Switzerland and the UK maintained conservative portfolios focused on operational optimisation. These differences highlight that innovation pathways are not linear but path-dependent and contextually embedded, advancing theoretical understandings of digital transformation as an institutionally negotiated rather than technologically determined pro.

### 4.3 Limitations

While this study provides a comparative and multidimensional analysis of home care business models and digital health adoption, it is important to critically acknowledge several limitations. These limitations arise both from the general research design and from the methods used to address each research question.

First, the reliance on the BMC as an analytical framework offered a useful structure for identifying and comparing business model elements across providers, but its applicability across diverse national contexts is limited. The BMC privileges commercial dimensions and may underrepresent relational, cultural, or institutional factors that strongly influence care provision. This limits the comprehensiveness of the findings and suggests that the framework should be treated as heuristic rather than definitive.

Second, the identification of leading providers was constrained by data availability. While the triangulated approach of revenue, client base, and geographic scope addressed some of the weaknesses of single-metric rankings, inconsistent disclosure of these indicators across countries reduced comparability. In fragmented systems or where small non-profits dominate, innovative providers may be systematically absent from datasets due to limited transparency or proprietary ownership structures. This introduces a selection bias that cannot be fully overcome within the current design.

Third, the analysis of business model patterns relied primarily on documentary evidence and expert interviews. While this combination allowed triangulation of perspectives, it remains vulnerable to reporting bias and self-presentation strategies by providers. Moreover, cross-national comparisons are necessarily limited by differences in institutional structures, policy environments, and terminologies. Readers should therefore treat the comparative insights as indicative patterns rather than definitive classifications.

Fourth, the mapping of DHTs was conducted using the Digital Therapeutics Alliance categorisation. While this provided an established taxonomy, it excluded non-health system software (e.g., HR or finance systems) and diagnostic technologies that may indirectly shape provider capacity and care quality. Moreover, the cross-sectional nature of the data limits insights into the temporal dynamics of adoption. Emerging technologies may not yet be reflected in provider portfolios, and observed gaps may therefore partly reflect timing rather than systemic neglect.

More broadly, the study’s country sample - guided by the GII (39) - prioritised contexts with broad innovation capacity. While this captured technologically advanced health systems, it may have overlooked countries with strong health-specific digital practices but lower general innovation scores. This introduces another potential bias and limits the generalisability of the findings to a broader global context.

### 4.4 Future Research

Future research can address these limitations in several ways. First, the development of validated and standardised outcome indicators - including revenue, client volumes, quality metrics, and workforce conditions - would enable more reliable cross-national comparisons. Longitudinal designs could further capture how providers adapt their business models in response to demographic pressures, regulatory reforms, and technological innovation.

Future studies should explore the real-world implementation of DHTs in greater depth. This includes barriers to adoption, interoperability challenges, workforce training, change management processes, and patient engagement. Understanding these dynamics will be critical for designing policies that support scalable innovation in home-based care.

Furthermore, further research should integrate broader indicators of performance, such as patient-reported outcomes, staff wellbeing, and equity measures, to complement financial and technological analyses. This would contribute to a more holistic understanding of what constitutes a high-performing home care model.

Comparative studies should examine the scalability and transferability of hybrid and socially embedded models. Attention should be paid to how institutional logics, financing structures, and cultural norms condition adoption across different health systems. Rather than assuming successful models can be replicated wholesale, future work should emphasise adaptive design and contextual fit, ensuring that business model innovations align with local institutional realities, workforce structures, and patient expectations.

## 5. Conclusion

This study provides a cross-national analysis of leading home care providers, highlighting how business model patterns and DHTs shape service delivery. While Fee-for-Service models remain dominant, we observed gradual movement toward value-based structures and revenue diversification. DHTs were mainly used for clinical documentation and operational efficiency, with only a subset of providers adopting advanced tools such as remote monitoring or digital therapeutics. Cross-country differences underscored the influence of local policies, financing, and cultural factors, showing that conventional welfare regime typologies alone cannot explain the diversity of models. Overall, the findings emphasize the need for context-sensitive innovation supported by regulatory flexibility, sustainable financing, and robust digital infrastructure to ensure scalable, digitally-enabled home care.

## Data Availability

All data produced in the present work are contained in the manuscript.

## List of abbreviation

AHP: Analytic Hierarchy Process
BMC: Business Model Canvas
DHTs: Digital Health Technologies
Digital: Therapeutics Digital Therapeutics
EMRs: Electronic Medical Records
GDP: Gross Domestic Product
GII: Global Innovation Index
KPIs: Key Performance Indicators
OECD: Organisation for Economic Co-operation and Development
VBP: Value-Based Payment
WHO: World Health Organization

## Declarations

### Ethics approval and consent to participate

This study received an ethics exemption from the Ethics Committee of the University of St. Gallen. The exemption was granted on the grounds that the research did not fall under the scope of the Swiss Federal Act on Research Involving Human Beings (Human Research Act, HRA), did not involve vulnerable populations, did not expose participants to physical or psychological risk, and did not require the collection of sensitive personal data.

### Consent for publication

All participants gave explicit consent for the use of anonymized quotations in publications resulting from this study.

### Availability of data and materials

The datasets generated and/or analysed during the current study are not publicly available due to participant confidentiality and data protection regulations but are available from the corresponding author on reasonable request.

### Authors’ contributions

PH, RV and TK designed the study. PH and SS conducted the interviews and jointly led the data analysis and manuscript writing. PH, RV and TK supervised the research process and provided critical feedback and revisions. All authors contributed to the interpretation of findings and approved the final manuscript.

## Acknowledgements

The authors would like to thank all study participants for their time and valuable insights.

## Appendices

## Appendix A. Interview Guide

### Customer Segments

- What different types of customers use your care services most frequently?
- How do you tailor your services to the needs of different customer groups?
- Are there any specific customer groups you specialize in?

### Value Proposition

- What specific services do you offer your customers?
- What are the key features of your service offering that set it apart from other providers?
- How do your services contribute to the improvement of your customers’ quality of life?

### Channels

- Through which channels do you reach your customers (e.g., in person, digital)?
- What role do physical vs. digital channels play in your customer communication?
- How do you use these channels to effectively convey your services and strengthen customer relationships?

### Customer Relationships

- How do you maintain long-term relationships with your customers?
- What measures do you take to ensure customer satisfaction and loyalty?
- Are there any special programs or initiatives to promote customer loyalty?

### Revenue Streams

- What models do you use to generate revenue (e.g., insurance billing, private payments)?
- How are your revenue streams developing in the context of changing healthcare regulations?
- Are there any seasonal fluctuations or trends that affect your revenue?

### Key Resources

- What resources are essential for the operation of your care services?
- How do you ensure the availability and quality of these resources?
- What role do technology and innovation play in your key resources?

### Key Activities

- What main activities are carried out daily to provide your services?
- What processes are crucial for the efficiency and quality of your services?
- How do you continuously optimize these processes?

### Key Partnerships

- With which partners do you collaborate to support your services (e.g., hospitals, care facilities, local authorities)?
- How do these partnerships contribute to strengthening your service offerings?
- Are there any particular challenges or benefits in these partnerships?

### Cost Structure

- What are the main cost factors for your care business?
- How do you manage these costs and strive for cost efficiency?
- Are there specific strategies to reduce costs without compromising service quality?

### Digitization and Technologies

- What digital tools and technologies do you use to improve your services?

- How do you integrate technological innovations into your daily operations and processes?
- Are there any specific digital initiatives or projects you are currently pursuing?
- What advantages and challenges do you see in the digitization of your services?

## Appendix B. Outreach Template

Dear [Provider Name],

I hope this message finds you well.

I am writing to you from the Center for Digital Health Interventions (CDHI), a collaborative initiative between ETH Zurich and the University of St. Gallen (HSG) in Switzerland. Our research at CDHI focuses on exploring the landscape of home care providers across various countries, with a keen interest in innovative technologies.

[Provider Name] has been identified as one of the leading providers in [Country], and we are keen to gain insights from your expertise. We would like to request a 30-minute interview with you to discuss your experiences and perspectives. In exchange, we are happy to share our research findings, as we believe in mutual learning and sharing among global leaders in the industry.

We greatly value your expertise and look forward to the possibility of collaborating on this important research project. Thank you for considering our request, and we appreciate your anticipated cooperation.

Best regards,

P.H.

## Appendix C. Definitions and Examples of Digital Health Technologies

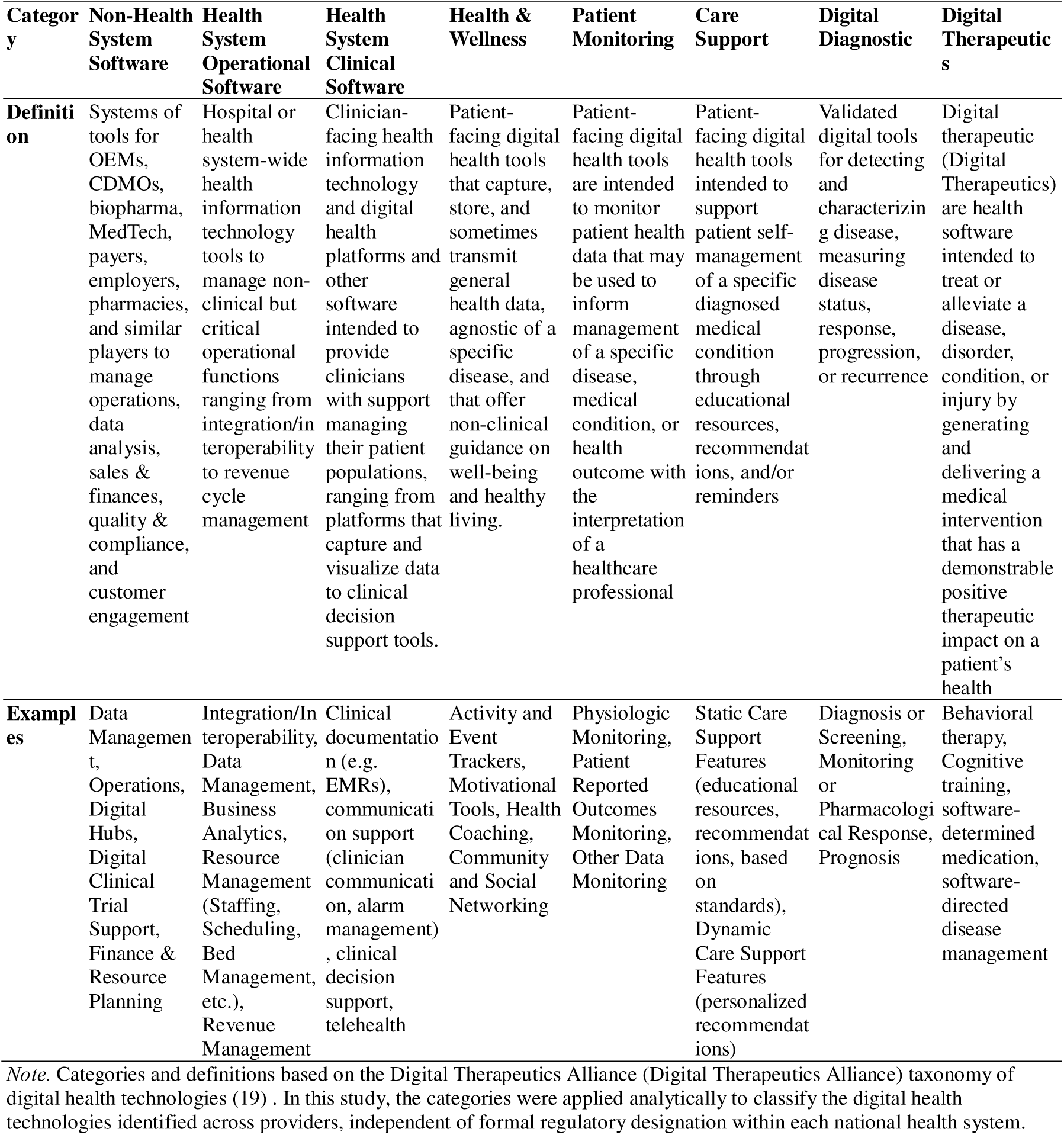

## Appendix D. Data Overview

| Provider ID | Providers | Country | Source I:<br>Service<br>Info | Source II:<br>Company<br>Info | Source III:<br>Market<br>Reports | Mode IV:<br>Interviews |
| --- | --- | --- | --- | --- | --- | --- |
| SG1 | All Saints Home | Singapore | 3 | 1 | 1 | 0 |
| SG2 | Home Nursing Foundation | Singapore | 3 | 2 | 1 | 0 |
| SG3 | St Luke's Eldercare | Singapore | 4 | 2 | 3 | 0 |
| SE1 | Attendo | Sweden | 4 | 2 | 2 | 1 |
| SE2 | Forenade Care | Sweden | 4 | 4 | 1 | 0 |
| SE3 | Vardaga (Ambea) | Sweden | 2 | 1 | 1 | 1 |
| CH1 | ASPMAD | Switzerland | 3 | 2 | 2 | 0 |
| CH2 | Spitex Zürich | Switzerland | 8 | 2 | 1 | 1 |
| CH3 | imad | Switzerland | 4 | 1 | 3 | 1 |
| UK1 | Bluebird Care | UK | 8 | 1 | 3 | 0 |
| UK2 | Helping Hands | UK | 5 | 1 | 3 | 0 |
| UK3 | Home Instead (UK) | UK | 17 | 1 | 4 | 2 |
| US1 | Amedisys | USA | 6 | 2 | 1 | 0 |
| US2 | Enhabit | USA | 9 | 3 | 3 | 0 |
| US3 | Maxim Healthcare | USA | 12 | 1 | 1 | 0 |
|  | **Total** | — | **92** | **26** | **30** | **6** |

*Note.* Five additional interviews were conducted with sector experts (e.g., consultants, policymakers, and association representatives). These interviews are not included above, as they were not linked to a specific provider.

## Appendix E. Business Models of Leading Home Care Providers

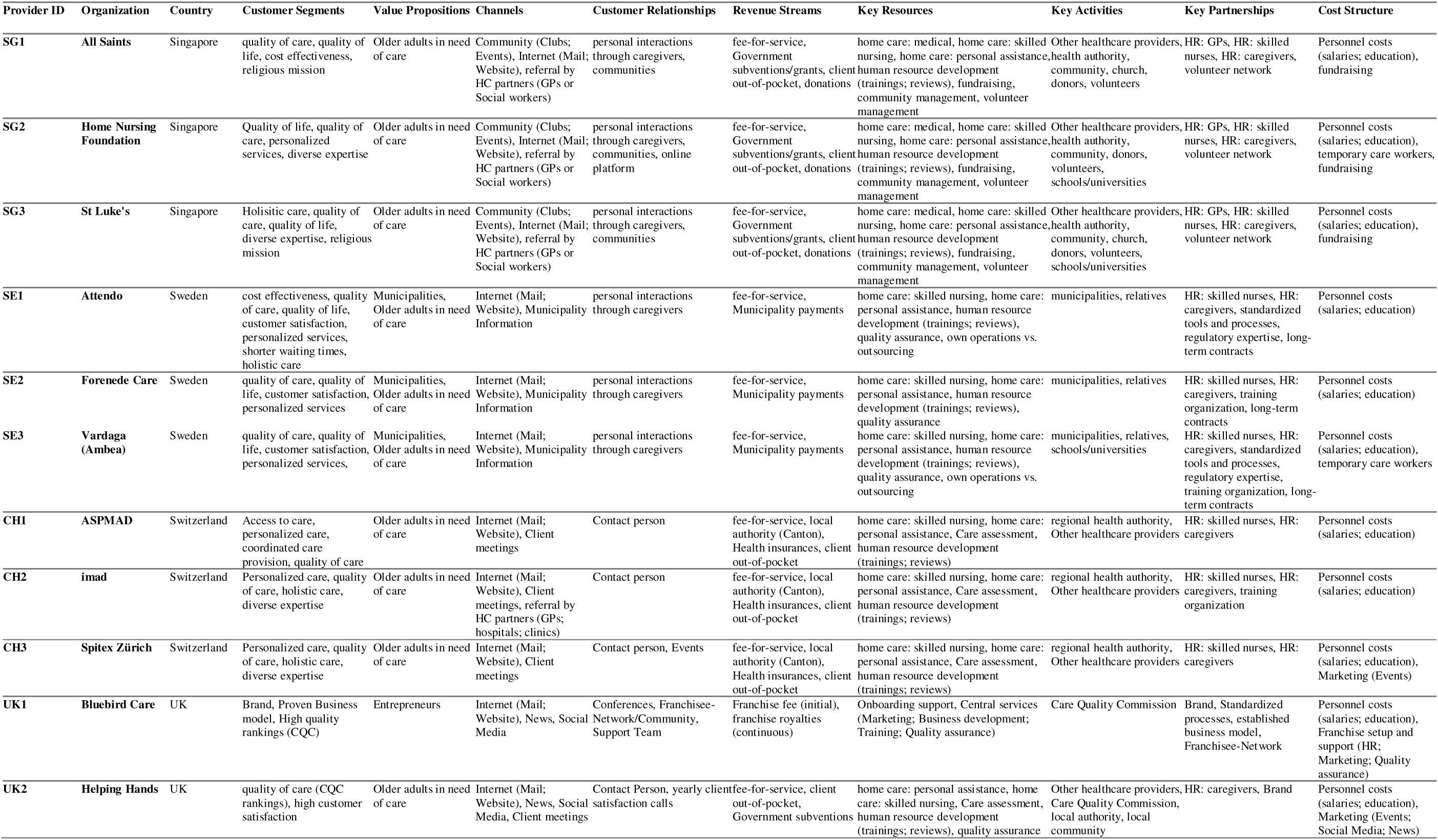
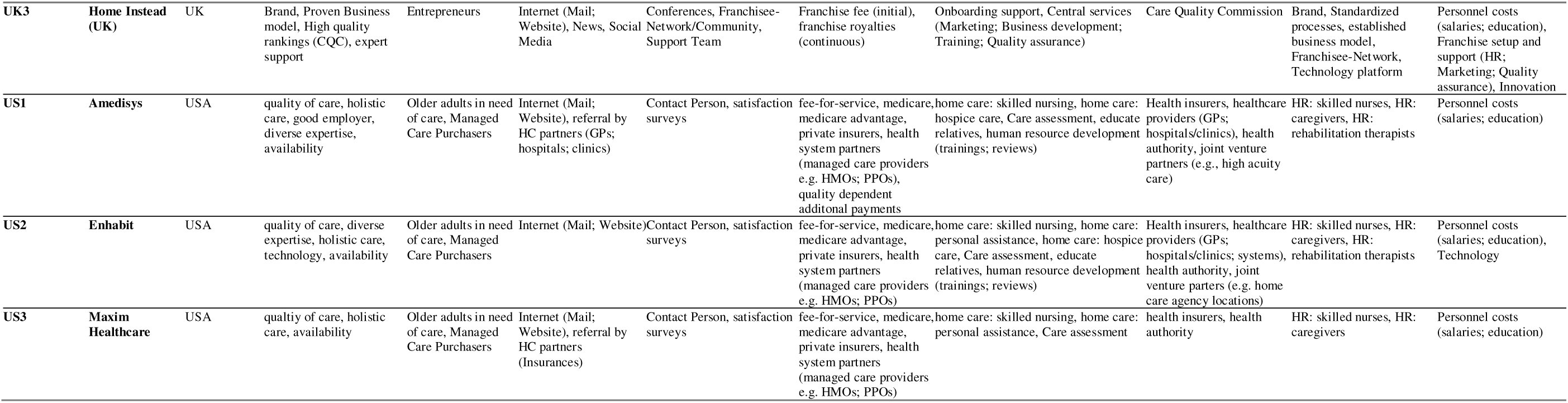

## Appendix F. Heatmap of Digital Health Technologies across Providers

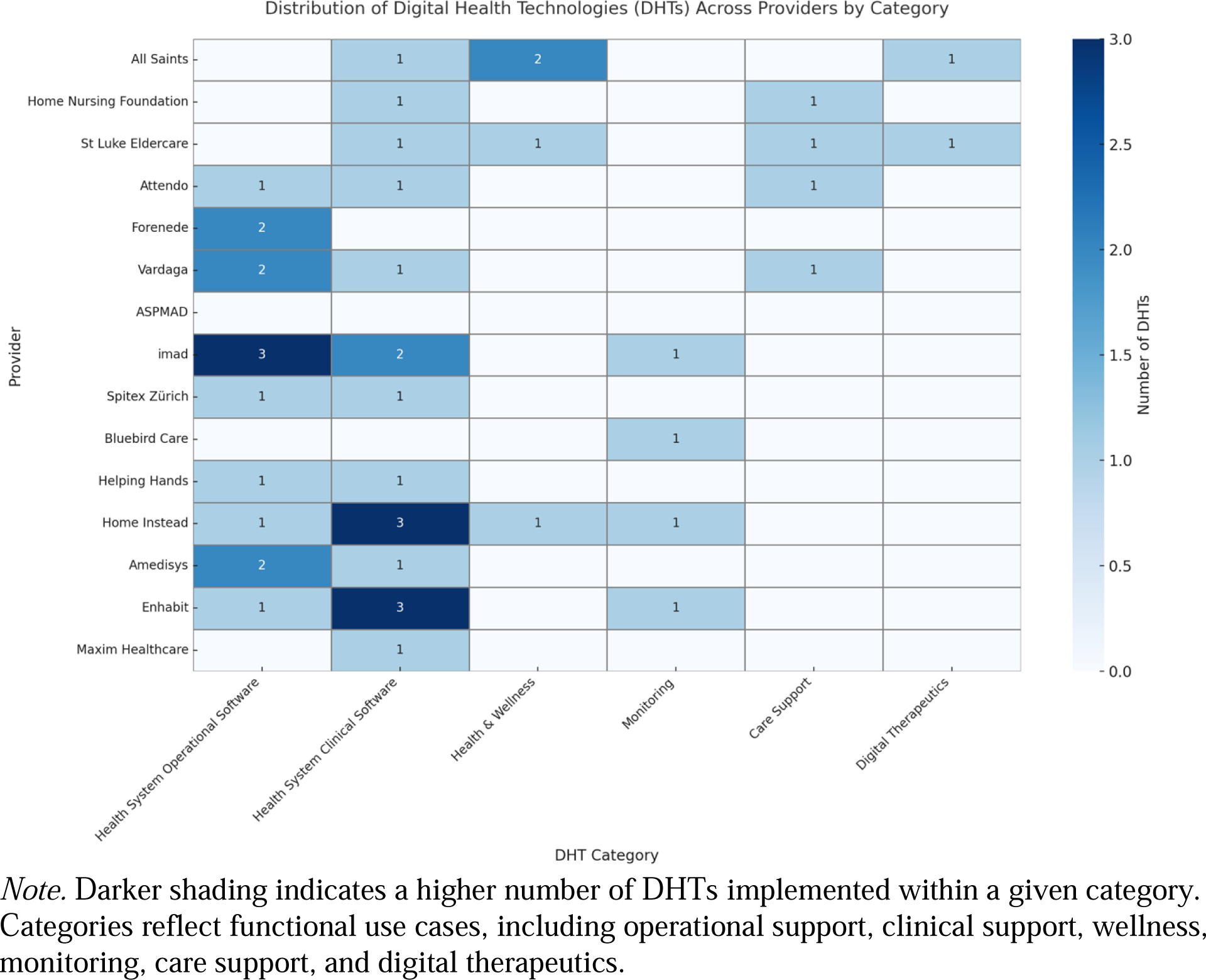

## Appendix G. Summarized Overview of Home Care Business Model Patterns Across Countries

|  | **Franchise Model** | **Fee-For-Service** | **Modification I** | **Modification II** | **Modification III** |
| --- | --- | --- | --- | --- | --- |
| **Country** | United Kingdom | Switzerland, Sweden, Singapore, US, United Kingdom | Sweden, United States, United Kingdom | Singapore | United States, Sweden |
| **Characteristics** | Standardized branding, centralized training, quality audits, shared digital infra, high regional penetration, locally adaptable service delivery | Direct service reimbursement, high labour costs, minute/hour-based billing, tariff regulation, care volume-driven revenue, staffing-intensive | Multi-sided market: B2C, B2G, B2B. Public/private payer mix. Contractual diversity to mitigate financial risk | Community involvement, philanthropic donations, volunteerism to fund supplementary care beyond reimbursements | Performance-linked payments, outcome-based incentives, quality metrics, EMR integration, care coordination improvements |

